# Inflammation biomarkers and neurobehavioral performance in rural adolescents

**DOI:** 10.1101/2024.10.15.24315322

**Authors:** Beemnet Amdemicael, Kun Yang, Briana N.C. Chronister, Caroline Mackey, Xin Tu, Sheila Gahagan, Danilo Martinez, Harvey Checkoway, David R Jacobs, Jose Suarez-Torres, Suzi Hong, Jose R. Suarez-Lopez

## Abstract

**Background:** Systemic inflammation has been associated with lower neurobehavioral performance in diverse populations, yet the evidence in adolescents remains lacking. Cytokines can alter neural network activity to induce neurocognitive changes. This work seeks to investigate the association between inflammation and neurobehavior in adolescents living in a rural region of Ecuador.

**Methods:** We examined 535 adolescents in rural communities of Ecuador (ESPINA study), 508 of which had neurobehavioral assessments (NEPSY-II) and circulating plasma levels of inflammatory markers (CRP, IL-6, TNF-⍺, sICAM-1, sVCAM-1, SAA, and sCD14). Associations between inflammatory biomarker concentrations and neurobehavioral scores were examined using adjusted bivariate semi-parametric models with generalized estimating equations. A partial least square regression approach was used to create composite variables from multiple inflammation biomarkers and model their association with cognitive outcomes.

**Results:** Higher sCD14 and TNF-α concentrations were significantly associated with lower social perception scores, by −0.47 units (95% CI: −0.80, −0.13) and −0.42 (−0.72, −0.12) for every 50% increase in inflammatory marker concentration, respectively. Similarly, every 50% increase in the inflammation summary score was associated with a significantly lower Social Perception score by −0.11 units (−0.19, −0.03). A unit increase in inflammatory composites of seven markers were associated with lower scores in language (−0.11 units, p=0.04), visuospatial processing (−0.15, p= 0.09), and social perception (−0.22, p=0.005) domains.

**Conclusions:** Higher levels of inflammation were associated with lower neurobehavioral performance in adolescents, especially with social perception. In addition, using a robust analytic method to examine an association between a composite inflammatory variable integrating seven markers led to additional findings, including the domains of language and visuospatial processing. A longitudinal follow-up of such investigations could unveil potential changes in inflammation-neurobehavior performance links through developmental stages and intervention opportunities.

## Introduction

Associations have been shown between greater levels of systemic inflammatory biomarkers and lower neurobehavioral performance or cognitive outcomes in diverse populations, including older adults with mild cognitive impairment and those with serious mental illness ^1–4^. In patients diagnosed with schizophrenia, increased blood levels of tumor necrosis factor alpha (TNF-α) were found to be significantly associated with reduced neurocognitive performances in processing speed, visual learning, and reasoning and problem solving ^5^. Similar findings in patients undergoing cardiopulmonary bypass further reinforce these findings, as an increase in C-reactive protein (CRP) and other proinflammatory cytokines were associated with neurocognitive decline in patients ^6^. Recently, inflammation predicted neurobehavioral performance in Corona Virus Disease 2019 (COVID-19) pneumonia survivors such that systemic inflammation, along with age, negatively correlated with performances in verbal memory, verbal fluency, speed of information processing and psychomotor coordination ^7^.

Understanding the underlying pathogenesis of the association between peripheral inflammation and cognition is of great interest to many investigators. Inflammatory cytokines, such as interleukin-1 (IL-1), interleukin-6 (IL-6) and tumor necrosis factor alpha (TNF-⍺) have been shown to alter the activity of neurotransmitter networks to induce neurobehavioral changes ^8^. Furthermore, elevated systemic levels of inflammatory biomarkers such as TNF-⍺, interleukins and interferons increase the expression of intracellular cell adhesion molecule 1 (ICAM-1) and vascular cell adhesion molecule 1 (VCAM-1) through upregulating gene transcription ^9^. ICAM-1, which is expressed on endothelial cells of the forebrain’s grey and white matter, as well as astrocytes and microglial cells, plays a crucial role in the blood brain barrier (BBB) ^10^. Proteolytic cleavage of ICAM-1 can transform the transmembrane glycoprotein into a soluble ICAM-1 (sICAM-1), whose levels have been associated with changes in permeability of the BBB, facilitating increased communication between the central nervous system (CNS) immune glial cells and the peripheral immune system ^10,11^. In an acute inflammatory state, ICAM-1 associated astrocytes and microglia, which heavily surround the capillary endothelial cells of the BBB, allow for the passage of peripheral immune cells, such as leukocytes, into the CNS parenchyma through astrocytic tight junctions and small vessel endothelia ^12^. This congruous process of increased peripheral and CNS immune activation has been shown to cause neuroinflammation and subsequent decline in neurocognition ^13^. Additionally, blood levels of immune activation markers such as soluble cluster of differentiation 14 (sCD14) have been found to increase monocytic inflammatory processes in the CNS implicated in cognitive declines ^14,15^. Though details of the underlying mechanisms have not been elucidated, high levels of plasma and cerebrospinal fluid sCD14 have been shown to have a significant association with markers of inflammation and axonal damage, further implicating sCD14 and monocytic activation in the pathogenesis of neurobehavioral decline ^16^. Notably, most findings characterizing this relationship between inflammation and neurobehavior have come from studies with predominantly adult populations and those with chronic pathology.

Despite the growing evidence in support of the association between inflammatory biomarkers levels and neurobehavior and cognitive outcomes, there are little data to substantiate the same association in adolescents. One of the few studies in young adults (18-24 years) included participants living with Human Immuno-Deficiency Virus (HIV) infection, among whom elevated plasma levels of sCD14, and sICAM-1 were negatively associated with visuospatial and verbal memory, respectively^17^. Adolescence is a period of important neurodevelopment during which even a subtle change in neurobehavioral outcomes may indicate potentially long-term impact. Thus, the aim of our study was to investigate the association between biomarkers of inflammation (CRP, IL-6, TNF-⍺, sICAM-1, sVCAM-1, serum amyloid A (SAA), and sCD14) and neurobehavioral performance in Ecuadorian adolescents. Based on prior findings, we hypothesize that higher levels of inflammation biomarkers will be associated with lower neurobehavioral performance.

## Methods

### Participants

The study of Secondary Exposures to Pesticides among Children and Adolescents (ESPINA - Exposición Secundaria a Plaguicidas en Niños y Adolescentes [Spanish]) is a prospective cohort focused on understanding the effects of pesticide exposure on the development of children and adolescents residing in the agricultural county of Pedro Moncayo, Pichincha Province, Ecuador. The ESPINA study was established in 2008 and examined 313 boys and girls of ages 4-9 years. Participants (73%) were recruited through community announcements for their participation in the “2004 Survey of Access and Demand of Health Services in Pedro Moncayo County,” a representative survey of Pedro Moncayo County collected by Fundación Cimas del Ecuador in conjunction with the communities of Pedro Moncayo County. Details about participant recruitment in 2008 have been described previously ^18^. In 2016 we expanded the cohort and conducted two examinations: in April, we examined 330 participants and in July-October we examined 535 participants for a combined total of 554 participants examined in 2016 (311 participants were examined at both time periods). Participants in 2016 were 12-17 years of age and included 238 participants examined in 2008 and 316 new volunteers. New participants in 2016 were invited to participate using the System of Local and Community Information (SILC) developed by Fundación Cimas del Ecuador, which included information of the 2016 Community Health Survey of Pedro Moncayo County (Formerly the Survey of Access and Demand of Health Services in Pedro Moncayo). In 2008 and 2016, none of the participants reported working in agriculture. Additional details of the participant recruitment in 2016 have been published ^19^.

To maximize the number of participants included in our analyses, we imputed missing data for parental education in 2016 (n=10), as well as residential distance to the nearest flower plantation in 2008 (n=3). Among the children who were examined in 2008 but were missing parental education data in 2016 (n=5), data for parental education were imputed using the 2008 data for maternal and paternal education. For participants with missing paternal and maternal education data in 2008 and missing parental education in 2016 (n=5), a random imputation was conducted for each variable based on a normal distribution of the variable during the respective examination period. Since a small number of participants reported being White (n=2), we grouped these 2 participants in the mestizo (mix of White and Indigenous) category to improve model stability when adjusting for race.

The 2016 examination of the ESPINA study was approved by the Institutional Review Boards of the University of California San Diego (UCSD), Universidad San Francisco de Quito and the Ministry of Public Health of Ecuador and endorsed by the Commonwealth of Rural Parishes of Pedro Moncayo County. We collected informed consent from adult participants (aged 18 years or older) and parents, as well as parental permission of participation and informed assent of child participants.

### Data Collection

#### Examination and Survey

In 2016, children were examined in 7 schools when they were out of session: first in April and again between July and October. The examiners were unaware of the participants’ potential exposures. Home interviews with parents and adult residents were conducted to gather socio-economic and demographic information.

#### Anthropometric measures

The height of the children was assessed using a height board following recommended guidelines, and the weight was measured using a digital scale (Tanita model 0108 MC; Tanita Corporation of America, Arlington Heights, IL, USA). We calculated z-scores for height-for-age (Z-height-for-age) and body mass index-for-age (Z-BMI-for-age) using the World Health Organization (WHO) growth standards ^20^.

#### Neurobehavioral Assessments

Neurobehavioral performance was measured using the NEPSY-II test (NCS Pearson, San Antonio, TX) ^21^. Neurobehavioral assessments were conducted in seven schools in Pedro Moncayo County. During the examinations of July-August 2008 and July-October 2016, trained psychologists blinded to participant exposure status assessed participants in 13 subtests across five domains: 1) Attention and Inhibitory Control (also known as Attention and Executive Functioning, subtests: Auditory Attention & Response Set, Inhibition); 2) Language (Comprehension of Instructions, Speeded Naming); 3) Memory and Learning (Immediate and Delayed Memory for Faces); 4) Visuospatial Processing (Design Copying, Geometric Puzzles); and 5) Social Perception (Affect Recognition). Two subtests required translation into Spanish using terminology appropriate for the local population (auditory attention and response set and comprehension of instructions). The translation was approved by NCS Pearson. Participants were examined alone and in a quiet room by the examiner. Participants who were also examined in the April 2016 examination completed 2 subtests at that time: Attention & Inhibitory Control and Inhibition. For this reason, it was necessary for us to adjust for learning effect associated with re-testing when analyzing these subtests (see Statistical Analysis).

We used the NEPSY-II scaled scores for each subtest to assess performance, which are age-standardized values based on a national sample of children in the United States ^22^. Scaled scores for the NEPSY-II subtest were calculated using the NEPSY-II scoring assistant software (NCS Pearson, Inc., San Antonio, TX) and higher scores indicate better performance. A detailed description of subtest scoring has been published elsewhere ^21,23,24^. Domain scores were used as measures of neurobehavioral performance and were calculated by averaging one primary scaled score from all subtests within each domain. For subtests that included either correct and error components (i.e. auditory attention and response set) or time and error components (i.e. inhibition, speeded naming, visuomotor precision), the combined scaled scores representing the combination of both components were used as primary scaled scores. Affect recognition was the only subtest in the Social Perception domain and, as such, the Social Perception domain is equivalent to the affect recognition scaled score. In this analysis, the sample sizes varied across domains because the NEPSY-II subtests were developed for different age ranges and were implemented only among participants in the appropriate age ranges.

### Biospecimen analysis

#### Plasma inflammatory markers

Venipuncture of the median cubital or cephalic veins (arm) were conducted by a phlebotomist following standard guidelines ^25^. Participants were not asked to fast prior to the venipuncture. Blood samples were centrifuged and aliquoted on site and transported on dry ice to Quito for storage at −80°C. Samples were then shipped to the University of California San Diego (UCSD) frozen using an overnight courier. Samples were stored at −80°C at UCSD until they were analyzed for inflammation marker quantification at the UCSD Integrative Health Mind-Body Biomarker Core. Multiplex assay kits were used to measure plasma levels of TNF-α and IL-6 to assess overall inflammatory states (*V-PLEX immunoassay proinflammatory kit*) and CRP, SAA, sICAM-1, and sVCAM-1to estimate vascular inflammatory states (*V-PLEX immunoassay vascular injury kit*), using the 2400 SECTOR Imager reader (Meso Scale Discovery, Rockville, Maryland). Circulating levels of cellular adhesion molecule can also reflect activation of endothelial cells, which can be implicated in impaired endothelium barrier functions including blood brain barrier^26^. Levels of sCD-14 were measured to assess monocyte activation which has been observed being associated with neuropsychological domains as described in the introduction, using quantikine ELISA kits using R& D Systems (Minneapolis, MN). The intra- and inter-assay coefficients of variation of those markers were below 10%.

Acetylcholinesterase (AChE) activity and hemoglobin concentration were measured in fresh finger-stick blood samples using the Environmental Quality Management (EQM)-Test-mate Cholinesterase (ChE) Test System 400 during the examination (EQM Research Inc., Cincinnati, OH, USA), and were collected in the April and July-October examinations. Lower concentrations of AChE activity reflect greater exposures to cholinesterase inhibitor pesticides ^27^.

The present analysis includes 508 participants examined in the July-October 2016, who had all covariates of interest: 8 observations were excluded due to missing neurobehavioral data and 19 due to missing covariate data.

### Statistical analysis

#### Participant characteristics

Descriptive sample characteristics include means and standard deviations (SD) for continuous variables and percent distributions for categorical variables, stratified by tertiles of the inflammation summary score (Table 1). The inflammation summary score was calculated by averaging the z-scores of all log-transformed inflammatory markers. We calculated the crude associations of the inflammation summary score and participant characteristics using linear regression for continuous characteristic variables and binomial regression for categorical characteristics.

**Table 1.** Participant characteristics.

|  | **Total**<br>**(N=508)** | **Inflammation Summary Score** |  |  | **P-Value** |
| --- | --- | --- | --- | --- | --- |
|  |  | **Q1**<br>**(N=169)** | **Q2**<br>**(N=169)** | **Q3**<br>**(N=170)** |  |
| **Age (years)** | 14.5 ± 1.75 | **14.9 ± 1.79** | **14.5 ± 1.71** | **14.1 ± 1.68** | **< 0.001** |
| **Gender, % Female** | 49% | 49% | 47% | 50% | 0.86 |
| **Race, % Mestizo** | 79% | 80% | 80% | 77% | 0.65 |
| **Height-for-Age**<br>**Z-Score, SD** | -1.48 ± 0.89 | -1.52 ± 0.82 | -1.48 ± 0.89 | -1.43 ± 0.97 | 0.40 |
| **Parental Education,**<br>**years** | 8.10 ± 3.49 | 8.03 ± 3.57 | 8.04 ± 3.62 | 8.24 ± 3.28 | 0.41 |
| **sCD14, ng/mL** | 1580 ± 385 | **1190 ± 147** | **1550 ± 92.3** | **2010 ± 273** | **< 0.01** |
| **CRP, µg/mL** | 2.69 ± 9.74 | **2.33 ± 9.75** | **2.64 ± 11.80** | **3.09 ± 7.08** | **< 0.01** |
| **IL-6, pg/mL** | 1.24 ± 1.42 | **1.07 ± 0.635** | **1.16 ± 0.939** | **1.49 ± 2.17** | **< 0.01** |
| **SAA, µg/mL** | 7.5 ± 40.6 | **4.40 ± 19.4** | **6.97 ± 43.7** | **11.1 ± 51.4** | **0.03** |
| **sICAM-1, ng/mL** | 758 ± 499 | 780 ± 519 | 726 ± 450 | 767 ± 528 | 0.57 |
| **sVCAM-1, ng/mL** | 973 ± 567 | 968 ± 516 | 958 ± 562 | 993 ± 620 | 0.86 |
| **TNF-α, pg/mL** | 2.11 ± 0.68 | **1.97 ± 0.52** | **2.08 ± 0.50** | **2.30 ± 0.91** | **< 0.01** |

Displayed values are mean ± standard deviation or percent.

#### Individual associations of inflammatory biomarkers with neurobehavior domains

Skewed variables, including the inflammatory biomarkers and inflammation summary score, were natural log-transformed ^28^, to fulfill the linear mixed model assumption criteria. Associations between the naturally log-transformed inflammatory biomarker concentrations and Inflammation summary score with neurobehavior assessment scores were assessed using adjusted bivariate semi-parametric model with a generalized estimating equation (GEE) model. The β estimates were multiplied by log(1.5) to describe the change of neurobehavioral assessment score per a 50% increase in the inflammatory biomarker concentration or inflammation summary score.

Covariates were included for the following potential confounders: age, gender, race, BMI-for-age z-score, height-for-age z-score, parental education level, AChE: hemoglobin ratio, creatinine, smokers at home (yes/no), and prior assessment (necessary only for Attention & Inhibitory Control Domain to control for test-retest performance differences, as a select number of participants were evaluated in this domain during both the April 2016 and July-October 2016 examinations). Covariates that were included in the final adjustment model were selected using backwards elimination and kept if they were statistically significant (p< 0.05). The following covariates were kept: age, race, height-for-age z-score, parental education level, and prior assessment the latter, only for the outcome of attention and inhibitory control. Although gender was not statistically significant, it was still included in the next iteration as this was a co-variate defined a-priori and neurobehavioral performance differences have been observed between boys and girls within the ESPINA study ^23^.

#### Inflammation mixture modeling and neurobehavioral performance

We created composite variables from multiple inflammation biomarkers to improve power when modeling their associations with cognitive outcomes. As the biomarkers serve as the independent variables in our analysis, we opted for the partial least squares (PLS) regression approach, rather than the principal component analysis (PCA) and factor analysis (FA). Like PCA and FA, PLS also create composite variables that are linear combinations of the original variables (biomarkers in the current context). However, unlike PCA and FA, PLS creates composite variables by also taking into account the relationship with the dependent variable. Specifically, the first composite has the maximum correlation with the dependent variable, followed by the second and so on. Hence, if interest lies in finding a subset of the original explanatory variables in the linear model that explains the most variability in the response as in our context, PLS composite variables are more effective than PCA or FA. The loadings of the composite variables represent the differential contributions by each independent variable, in this case each inflammation biomarker. A positive direction of the loading of a biomarker indicates that the biomarker has the same direction as the composite when interpreting the association of the biomarker with the dependent outcome in the regression analysis (see Figure 1a). In addition to p-values, the effect size, adjusted R-squared, is also used to determine if it is important to include a composite variable in the regression model, which is especially helpful if the effect size of the composite is not large (Figure 1). The inflammation biomarker concentrations were normally standardized for PLS regression to eliminate the scale differences between them. Particularly for this study, a residualization procedure was adopted as adjustment for covariates, which means the outcome variable corresponding to each cognitive outcome was the residual we got from a linear model fitted with the original outcome and the covariates. All analyses were conducted using R Version 4.2.2

**Figure 1.**
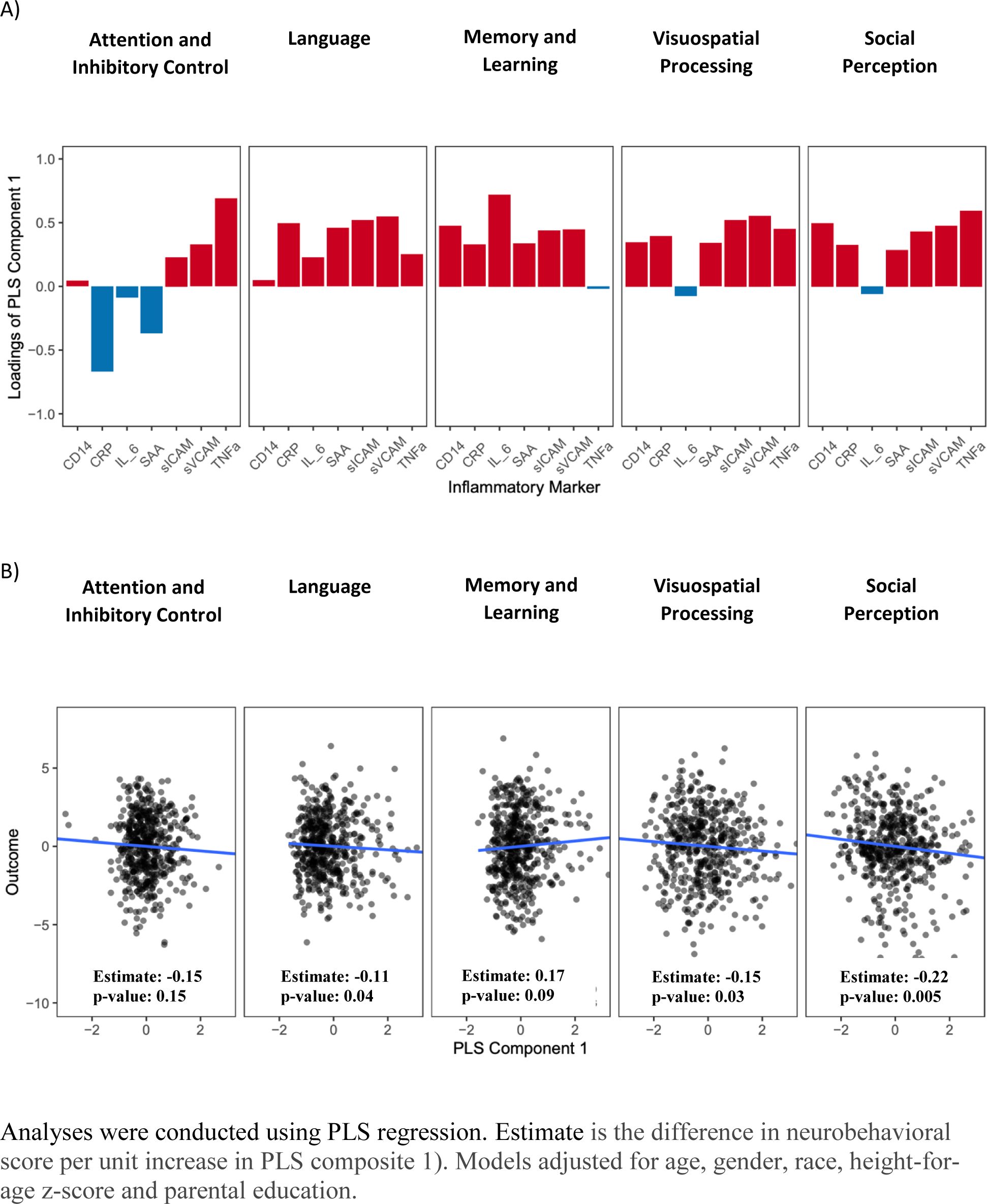
A) Loadings of the PLS Inflammation Component 1; B) associations of PLS Inflammation Component 1 with neurobehavioral domain scores in adolescent participants of the ESPINA study. N=508.

## Results

### Participant characteristics

The mean (SD) age of participants was 14.5 years (1.75), and 49% of the participants were female, with 79% of participants being Mestizo. The mean (SD) years of parental education was 8.10 years (3.49), and participants were on average 1.48 SD (0.89) shorter (height-for-age z-score) than the WHO normative sample. The average participant scores in all neurobehavioral domains were lower than the NEPSY-II normative sample mean score of 10 (Table S1). Age was negatively associated with the inflammation summary score (p<0.001, Table 1). Levels of all inflammatory biomarkers were significantly higher in the higher inflammation summary score groups (p<0.001) as expected, except for sICAM-1 and sVCAM-1.

### Independent associations between inflammatory markers and neurobehavior domains

Higher sCD14 and TNF-α concentrations were significantly associated with lower social perception scores, by −0.465 units (95% CI: −0.80, −0.13) and −0.418 units (−0.72, −0.12) for every 50% increase in inflammatory marker concentration, respectively (Table 2). Similarly, every 50% increase in the inflammation summary score was associated with a significantly lower Social Perception score by −0.112 units (−0.19, −0.03) and a borderline non-significant association Visuospatial Processing score by −0.054 units (−0.13, 0.02). No other significant associations were found between the remaining five inflammatory biomarkers and neurobehavioral domain scores. Yet, most inflammation-neurobehavior associations shown in Table 2 were also negative for the domains of Language, Memory & Learning, Visuospatial Processing and Social Perception. We did not observe evidence of effect modification by gender or age.

**Table 2.** Associations between inflammatory biomarker level and neurobehavioral performance in adolescents of the ESPINA study. N= 508.

|  | Neurobehavioral Domain |  |  |  |  |
| --- | --- | --- | --- | --- | --- |
|  | Attention &<br>Inhibitory<br>Control | Language | Memory &<br>Learning | Visuospatial<br>Processing | Social<br>Perception |
| **sCD14** | 0.01<br>(-0.30, 0.32) | 0.06<br>(-0.23, 0.35) | 0.17<br>(-0.17, 0.50) | -0.22<br>(-0.52, 0.08) | **-0.47**<br>**(-0.80, -0.13)** |
| **CRP** | -0.01<br>(-0.06, 0.05) | 0.01<br>(-0.04, 0.06) | 0.03<br>(-0.03, 0.08) | -0.03<br>(-0.09, 0.02) | 0.00<br>(-0.06, 0.06) |
| **IL-6** | -0.01<br>(-0.15, 0.13) | -0.12<br>(-0.24, 0.01) | -0.04<br>(-0.19, 0.12) | 0.00<br>(-0.16, 0.16) | -0.10<br>(-0.27, 0.07) |
| **SAA** | -0.01<br>(-0.06, 0.04) | -0.01<br>(-0.07, 0.05) | 0.02<br>(-0.04, 0.08) | -0.04<br>(-0.10, 0.02) | 0.01<br>(-0.06, 0.08) |
| **sICAM-1** | -0.00<br>(-0.13, 0.12) | 0.01<br>(-0.12, 0.15) | 0.02<br>(-0.13, 0.17) | -0.06<br>(-0.21, 0.08) | -0.03<br>(-0.18, 0.12) |
| **sVCAM-1** | -0.00<br>(-0.14, 0.13) | -0.03<br>(-0.17, 0.12) | -0.01<br>(-0.17, 0.14) | -0.08<br>(-0.24, 0.08) | -0.09<br>(-0.26, 0.07) |
| **TNFα** | 0.01<br>(-0.26, 0.28) | -0.10<br>(-0.38, 0.19) | -0.09<br>(-0.39, 0.20) | -0.09<br>(-0.44, 0.26) | **-0.42**<br>**(-0.72, -0.12)** |
| **Inflammation**<br>**summary score** | 0.00<br>(-0.07, 0.08) | 0.02<br>(-0.06, 0.09) | 0.04<br>(-0.04, 0.12) | -0.05<br>(-0.13, 0.02) | **-0.11**<br>**(-0.19, -0.03)** |

Analyses were conducted using GEE models. Displayed values are score differences in NEPSY-II scores in relation to a 50% increase in inflammatory biomarker level (95% confidence interval).
Models adjusted for age, gender, race, height-for-age z-score, and parental education. Associations with Attention & Inhibitory Control were also adjusted for recent prior assessment in April 2016 (test-retest). Bolded text indicates statistical significance for the 95% confidence interval.

### Composite inflammatory marker associations with neurobehavior domains using PLS regression

We applied PLS to construct inflammation composite variables from all available inflammatory biomarkers (sCD14, CRP, IL-6, SAA, sICAM-1, sVCAM-1 and TNF-α), and modeled the associations of extracted composite variables with neurobehavioral domains using linear regression, controlling for covariates. Our final models include only the first PLS composite variable, as sequentially added second and third composites resulted in minimal improvements in the adjusted R-square for the model compared to the first composite variable only (Table 3).

**Table 3.** Associations between PLS Inflammation Components and Neurobehavioral Performance in adolescent participants of the ESPINA study. N= 508.

| Neurobehavioral Domain | Total $R^2_{adj}$ | PLS Inflammation Component | | |
| --- | --- | --- | --- | --- |
|  |  | 1 | 2 | 3 |
| Attention & Inhibitory Control | -0.0081 | -0.15,<br>95% CI:<br>[-0.35, 0.05],<br>$R^2_{adj} = 0.002$ | -0.09,<br>95% CI:<br>[-0.32, 0.14],<br>$R^2_{adj} = 0.001$ | -0.02,<br>95% CI:<br>[-0.15, 0.12],<br>$R^2_{adj} = -0.001$ |
| Language | 0.0005 | **-0.11,**<br>**95% CI:**<br>**[-0.22, -0.004],**<br>**$R^2_{adj} = 0.006$** | -0.105,<br>95% CI:<br>[-0.28, 0.07],<br>$R^2_{adj} = 0.007$ | -0.18,<br>95% CI:<br>[-0.52, 0.15],<br>$R^2_{adj} = 0.007$ |
| Memory & Learning | -0.0060 | -0.17,<br>95% CI:<br>[-0.36, 0.02],<br>$R^2_{adj} = 0.004$ | -0.05,<br>95% CI:<br>[-0.19, 0.09],<br>$R^2_{adj} = 0.003$ | -0.05,<br>95% CI:<br>[-0.28, 0.17],<br>$R^2_{adj} = 0.001$ |
| Visuospatial Processing | -0.0026 | **-0.15,**<br>**95% CI:**<br>**[-0.29, -0.01],**<br>**$R^2_{adj} = 0.007$** | -0.08,<br>95% CI:<br>[-0.23, 0.08],<br>$R^2_{adj} = 0.007$ | -0.04,<br>95% CI:<br>[-0.33, 0.24],<br>$R^2_{adj} = 0.005$ |
| Social perception | 0.0052 | **-0.22,**<br>**95% CI:**<br>**[-0.38, -0.07],**<br>**$R^2_{adj} = 0.013$** | -0.08,<br>95% CI:<br>[-0.22, 0.06],<br>$R^2_{adj} = 0.014$ | -0.10,<br>95% CI:<br>[-0.49, 0.28],<br>$R^2_{adj} = 0.012$ |

Analyses were conducted using PLS regression. Displayed values are $\beta$ coefficient (difference in neurobehavioral score per unit increase in PLS composite 1). The outcomes (neurobehavioral scores) are residualized by the following covariates: age, gender, race, height-for-age z-score, and parental education. Estimates corresponding to the 3 components are calculated with additive models respectively (PLS 1, PLS1+2, PLS1+2+3) Bold indicates statistical significance by the 95% confidence interval.

As expected, the loadings of each inflammation marker on each of the components differed across neurobehavioral outcomes. First, shown in Figure 1a are loadings of each inflammatory marker on the first inflammation PLS composite variable; loadings shown in red indicate that the biomarker contributed positively to the first PLS composite variable, whereas loadings shown in blue indicate negative contributions. Furthermore, as shown in Figure 1b, the first inflammation PLS component was negatively associated with language (β coefficient [difference in neurobehavioral score per unit increase in inflammation PLS composite 1]= −0.11, p=0.043), visuospatial processing (β= −0.15, p= 0.086), and social perception (β= −0.22, p=0.005; Figure 1b, Table 3) domains. As indicated by the positive loadings of inflammatory markers on PLS component 1, a greater inflammatory state, as reflected by greater levels of sCD14, CRP, SAA, sICAM-1, sVCAM-1 and TNF-α, was associated with lower scores in Language, Visuospatial Processing and Social Perception in this cohort of adolescents. Only IL-6 showed negative loadings on the first PLS components but to a minimal degree and did not contribute to neurobehavioral performance outcomes.

## Discussion

Our study revealed certain associations of interest between circulating inflammatory molecules and neurobehavioral performance in an Ecuadorian adolescent population. As hypothesized, greater levels of circulating inflammatory markers were associated with lower neurobehavioral performance. A series of regression analyses assessing independent associations for each inflammatory biomarker led to the finding that an overall inflammation score, sCD14 and TNF-α concentrations were negatively associated with scores of Social Perception domain. This finding was replicated in a more integrated examination using PLS regression, in which the effect of a composite of inflammatory markers was investigated. The PLS regression approach yielded findings in which higher composites of various inflammatory markers (PLS component 1) were associated with lower Language, Visuospatial Processing, and Social Perception scores. The most consistent contributors to the PLS component 1 for each of the neurobehavioral domains were sICAM-1 and sVCAM-1, followed by CRP, SAA and TNFα. In addition, TNFα and sCD14 were important contributors to PLS component 1 in its association with visuospatial processing and social perception, suggesting a potential role of monocytes in these associations as well as hepatocytes as another source of sCD14 in circulation^29^. Our findings suggest that a more comprehensive and integrated approach that considers multiple inflammatory markers is a more robust approach to scrutinize the link between inflammation and neurocognition.

Our findings agree with those from prior studies showing inverse associations of sCD14 with neurocognitive outcomes. Among individuals living with HIV who experienced neurocognitive decline, peripheral immune activation characterized by increased blood levels of sCD14 and activation of glial cells resulting in neuroinflammation and neuronal injury, were correlated with CNS inflammation and HIV-associated neurocognitive disorders ^30,31^. Higher plasma levels of sCD14 were also associated with a global decline in the domains of attention and learning in participants with and without HIV associated neurocognitive disorder, including visuospatial memory ^15, 17^. Of note, the average circulating levels of sCD14 found in our study were comparable to those (1468.04 ± 420.66 ng/mL) reported in young adults living with HIV and treated with antiretroviral treatment reported in the study by Kim-Chang and colleagues ^17^. Similarly, a study from East Africa report chronic inflammation and elevated sCD-14 levels were associated with lower neurobehavioral performance in participants, regardless of HIV status^32^. These findings of the sCD14-cogntion association indicate an important role of monocytic inflammatory processes in the CNS outcomes ^15^. More specifically, the cortical and limbic pathways, home to an abundance of chemoattractants with a preference for monocytic recruitment, are thought to be involved in this pathophysiology. ^15,33,34^. Once recruited to the CNS, activated monocytes/macrophages contribute to a neuroinflammatory state and altered neurobehavior as indicated by the association between greater cerebrospinal fluid levels of sCD14 and impaired neurobehavioral performance ^14^. To our knowledge, there is paucity of evidence specifically indicating monocyte activation or inflammation underlying social perception, especially among adolescents. Given that social perception is a critical function from a neurodevelopmental perspective, the role of monocyte activation and validation of its markers is of significance (Pelphrey & Carter 2008).

Our results also show associations between other inflammatory markers and neurobehavioral functions, including language and visuospatial processing. These findings are corroborated in a population based Finish prospective cohort study, which found that elevated TNF-α and IL-6 independently predicted poorer neurocognition, specifically in verbal fluency and word list learning, suggesting they are frequent predictors of cognitive decline^2^. Within the CNS, cytokines released by activated microglia in response to elevated systemic inflammation can bind neuronal receptors and lead to changes in cognition, as well as mood and behavior ^36^. Inflammatory molecules such as TNF-α and CRP, play an important role in maintaining an equilibrium between synaptogenesis and neurodegeneration, which is crucial for cognition ^5,37^. Inflammatory disturbance of this equilibrium and subsequent axonal CNS injury have been proposed as a likely mechanism of neurocognitive deficit ^6,37^.

Notably, the impact of vascular endothelial inflammation should be highlighted in cognitive and behavioral outcomes. In adult populations, studies have shown that increased levels of circulating cellular adhesion molecules, such as sICAM-1, have been associated with lower cognitive function, while sVCAM-1 has been associated with older age and inflammatory microglial activation leading to diminished cognition and neurogenesis ^38–40^. Our findings in adolescents are in agreement with and extend beyond the existing literature in adult populations. Increased sICAM-1 and sVCAM-1 levels are indicative of vascular endothelial dysfunction ^41^. Endothelial injury and repair are affected by age such that endothelial injury increases with advancing age, while endothelial repair decreases with age as shown in a study including healthy children, adults, and children with familial hypercholesterolemia ^42^. The findings of our study are consistent with the aforementioned studies, despite participants’ young age and lack of vascular endothelial pathology.

In children with pulmonary arterial hypertension, pressure-induced endothelial damage was associated with activation of endothelial cells and subsequent increase in expression of cell adhesion molecules sICAM-1 ^43^. This may be a contributing pathophysiological pathway behind lower neurobehavioral scores observed among children with primary hypertension compared to normotensive counterparts ^44,45^. An endothelial damage-associated increase in soluble CAMs was associated with worse short-term memory, speed of processing, and executive function in adults ^40^. Furthermore, elevated levels of sVCAM-1 were found to be associated with increased cerebrovascular resistance and higher levels of cognitive impairment in verbal memory in elderly participants ^46^. Similarly, increased sVCAM-1 levels were found to be associated with worse cognition in patients with primary progressive multiple sclerosis and the odds of cognitive decline increased by 30% with each 100 ng/mL increase of sVCAM-1^47^.A separate study in elderly adults report that high levels of sICAM-1 are independently associated with greater cognitive impairment ^48^. While these findings are consistent with ours, it’s important to recognize that they were in adults of mid to older age. It is probable that vascular inflammation affects differing neurocognitive subdomains across the life course, and more studies are needed from childhood to young adulthood of diverse sociodemographic, geographic, culture, and environmental background.

Our study includes strengths such as a large sample size and inclusion of under-represented adolescent individuals, and multidimensional neurobehavior examination. Additionally, this is a population-based study that includes mostly healthy participants, which increases the generalizability of the study findings to other adolescent populations. Also, participants were examined during a relatively short amount of time (between July-October 2016), which reduces the role of seasonal variations on the independent and dependent variables evaluated. Meanwhile, some limitations are noted such as inflammatory markers that were assessed only in blood, leading to a challenge in deciphering mechanistic pathways. At the same time, reliable biomarkers that can be measured in blood with relative ease, representing consistent implications for functional outcomes have high utility. Given the link between peripheral and neuro-inflammation as aforementioned, concurrent examinations of markers of inflammation in blood and CNS would offer more direct mechanistic insight, although currently available methods to assess neuroinflammation in vivo such as positron emission tomography (PET) imaging are often costly and inaccessible. There remain needs for non-invasive and cost-effective methods to assess markers of neuroinflammation and interrogate the peripheral and CNS inflammation interactions. Additionally, our findings are based on a cross-sectional examination which limits the interpretation of the directionality of the associations and its developmental impact. Future longitudinal studies that enable examinations of inflammation and neurocognition from childhood to adulthood will offer valuable knowledge and inform prevention and intervention strategies as necessary.

## Conclusion

Our study provides novel insights into the associations between inflammatory markers and neurobehavioral performance in a population that is under-represented in this area of research. We found that higher levels of inflammation were associated with lower neurobehavioral performance in adolescents residing in a rural region of Ecuador. Independent associations of TNF-α and sCD14 with lower scores in social perception were found. An integrated examination incorporating all inflammation biomarkers using a PLS regression approach led to a finding that an overall greater systemic inflammatory state is associated with lower performance in the domains of language, visuospatial processing, and social perception in adolescents. Our findings also point to a few potentially promising vascular inflammatory markers in relation to those domains, although a closer examination in a longitudinal study is warranted. These are among the first findings to characterize these associations in adolescents, which help to elucidate the intricate relationship between inflammation and neurocognition during a neurodevelopmental period.

## Data Availability

All data produced in the present study are available upon reasonable request to the authors

https://knit.ucsd.edu/espina/

### Key Points

- Studies in adults have shown that greater levels of systemic inflammation are associated with lower neurobehavioral performance. There are little data to substantiate this association in adolescents.
- Higher levels of inflammation were associated with lower neurobehavioral performance in adolescents.
- TNF-α and sCD14 were independently associated with lower scores in social perception.
- A composite of inflammation (component 1) was statistically significantly associated with lower scores in the domains of Language, Visuospatial Processing and Social Perception.

### Relevance

- These findings make a valuable contribution to the sparse body of data within the realm of research focused on Ecuadorian adolescents. As the literature and evidence grows, we hope the newly uncovered knowledge can inform prevention and intervention policies as necessary.

## Acknowledgements

Research reported in this publication was supported by the National Institute of Environmental Health Sciences of the National Institutes of Health under Award Numbers (R01ES025792, R01ES030378, R21ES026084). B.N.C. Chronister was funded by the Institute of Mental Health (5T32MH122376). We thank ESPINA study staff, Fundación Cimas del Ecuador, the Parish Governments of Pedro Moncayo County, community members of Pedro Moncayo and the Education District of Pichincha-Cayambe-Pedro Moncayo counties for their contributions and/or support on this project.

## Abbreviations

CRP: C-Reactive Protein
IL-6: Interleukin-6
TNF-⍺: Tumor Necrosis Factor Alpha
sICAM-1: soluble Intracellular Cell Adhesion Molecule-1
sVCAM-1: soluble Vascular Cell Adhesion Molecule-1
SAA: Serum Amyloid A
CD14: Cluster of Differentiation 14

## References

1. Hope S, Hoseth E, Dieset I, et al. Inflammatory markers are associated with general cognitive abilities in schizophrenia and bipolar disorder patients and healthy controls. Schizophr Res. 2015;165(2):188–194. doi:10.1016/j.schres.2015.04.004

2. Kipinoinen T, Toppala S, Rinne JO, Viitanen MH, Jula AM, Ekblad LL. Association of Midlife Inflammatory Markers With Cognitive Performance at 10-Year Follow-up. Neurology. 2022;99(20):e2294–e2302. doi:10.1212/WNL.0000000000201116

3. Shen XN, Niu LD, Wang YJ, et al. Inflammatory markers in Alzheimer’s disease and mild cognitive impairment: a meta-analysis and systematic review of 170 studies. J Neurol Neurosurg Psychiatry. 2019;90(5):590–598. doi:10.1136/jnnp-2018-319148

4. Zheng M, Chang B, Tian L, et al. Relationship between inflammatory markers and mild cognitive impairment in Chinese patients with type 2 diabetes: a case-control study. BMC Endocr Disord. 2019;19(1):73. doi:10.1186/s12902-019-0402-3

5. Kogan S, Ospina LH, Kimhy D. Inflammation in Individuals with Schizophrenia – Implications for Neurocognition and Daily Function. Brain Behav Immun. 2018;74:296–299. doi:10.1016/j.bbi.2018.09.016

6. Ramlawi B, Rudolph JL, Mieno S, et al. C-Reactive protein and inflammatory response associated to neurocognitive decline following cardiac surgery. Surgery. 2006;140(2):221–226. doi:10.1016/j.surg.2006.03.007

7. Mazza MG, Palladini M, De Lorenzo R, et al. Persistent psychopathology and neurocognitive impairment in COVID-19 survivors: Effect of inflammatory biomarkers at three-month follow-up. Brain Behav Immun. 2021;94:138–147. doi:10.1016/j.bbi.2021.02.021

8. Berthold-Losleben M, Himmerich H. The TNF-α System: Functional Aspects in Depression, Narcolepsy and Psychopharmacology. Curr Neuropharmacol. 2008;6(3):193–202. doi:10.2174/157015908785777238

9. Dietrich JB. The adhesion molecule ICAM-1 and its regulation in relation with the blood– brain barrier. J Neuroimmunol. 2002;128(1):58–68. doi:10.1016/S0165-5728(02)00114-5

10. Müller N. The Role of Intercellular Adhesion Molecule-1 in the Pathogenesis of Psychiatric Disorders. Front Pharmacol. 2019;10. Accessed October 9, 2022. https://www.frontiersin.org/articles/10.3389/fphar.2019.01251

11. Ramos TN, Bullard DC, Barnum SR. ICAM-1: Isoforms and Phenotypes. J Immunol. 2014;192(10):4469–4474. doi:10.4049/jimmunol.1400135

12. Benveniste EN. Inflammatory cytokines within the central nervous system: sources, function, and mechanism of action. Am J Physiol-Cell Physiol. 1992;263(1):C1–C16. doi:10.1152/ajpcell.1992.263.1.C1

13. Subramaniyan S, Terrando N. Neuroinflammation and Perioperative Neurocognitive Disorders. Anesth Analg. 2019;128(4):781. doi:10.1213/ANE.0000000000004053

14. Kamat A, Lyons JL, Misra V, et al. Monocyte activation markers in cerebrospinal fluid associated with impaired neurocognitive testing in advanced HIV infection. J Acquir Immune Defic Syndr 1999. 2012;60(3):234–243. doi:10.1097/QAI.0b013e318256f3bc

15. Lyons JL, Uno H, Ancuta P, et al. Plasma sCD14 is a biomarker associated with impaired neurocognitive test performance in attention and learning domains in HIV infection. J Acquir Immune Defic Syndr 1999. 2011;57(5):371–379. doi:10.1097/QAI.0b013e3182237e54

16. Jespersen S, Pedersen KK, Anesten B, et al. Soluble CD14 in cerebrospinal fluid is associated with markers of inflammation and axonal damage in untreated HIV-infected patients: a retrospective cross-sectional study. BMC Infect Dis. 2016;16(1):176. doi:10.1186/s12879-016-1510-6

17. Kim-Chang JJ, Donovan K, Loop MS, et al. Higher soluble CD14 levels are associated with lower visuospatial memory performance in Youth with HIV (YWH). AIDS Lond Engl. 2019;33(15):2363–2374. doi:10.1097/QAD.0000000000002371

18. Suarez-Lopez JR, Jacobs DR, Himes JH, Alexander BH, Lazovich D, Gunnar M. Lower acetylcholinesterase activity among children living with flower plantation workers. Environ Res. 2012;114:53–59. doi:10.1016/j.envres.2012.01.007

19. Espinosa da Silva C, Gahagan S, Suarez-Torres J, Lopez-Paredes D, Checkoway H, Suarez-Lopez JR. Time after a peak-pesticide use period and neurobehavior among ecuadorian children and adolescents: The ESPINA study. Environ Res. 2022;204(Pt C):112325. doi:10.1016/J.ENVRES.2021.112325

20. World Health Organization. World Health Organization Child Growth Standards : Length/Height-for-Age, Weight-for-Age, Weight-for-Length, Weight-for-Height and Body Mass Index-for-Age : Methods and Development. World Health Organization, Department of Nutrition for Health and Development; 2006.

21. Kemp SL, Korkman M. Essentials of NEPSY-II Assessment. John Wiley & Sons; 2010.

22. Korkman M, Kirk U, Kemp SL. NEPSY II: Clinical and Interpretive Manual. 2nd ed. The Psychological Corporation; 2007.

23. Suarez-Lopez JR, Himes JH, Jacobs DR, Alexander BH, Gunnar MR. Acetylcholinesterase activity and neurodevelopment in boys and girls. PEDIATRICS. 2013;132(6):e1649–e1658. doi:10.1542/peds.2013-0108

24. Suarez-Lopez JR, Checkoway H, Jacobs DRJ, et al. Potential short-term neurobehavioral alterations in children associated with a peak pesticide spray season: The Mother’s Day flower harvest in Ecuador. Neurotoxicology. 2017;60(5):125–133. doi:10.1016/j.neuro.2017.02.002

25. Clinical and Laboratory Standards Institute. Procedures for the Collection of Diagnostic Blood Specimens by Venipuncture: Approved Standard. Sixth. (Ernst D, ed.). Clinical and Laboratory Standards Institute; 2007.

26. Sheikh MH, Henson SM, Loiola RA, et al. Immuno-metabolic impact of the multiple sclerosis patients’ sera on endothelial cells of the blood-brain barrier. J Neuroinflammation. 2020;17(1):153. doi:10.1186/s12974-020-01810-8

27. Skomal AE, Zhang J, Yang K, et al. Concurrent urinary organophosphate metabolites and acetylcholinesterase activity in Ecuadorian adolescents. Environ Res. 2022;207:112163. doi:10.1016/j.envres.2021.112163

28. Hair JF, Risher JJ, Sarstedt M, Ringle CM. When to use and how to report the results of PLS-SEM. Published online January 14, 2019. doi:10.1108/ebr-11-2018-0203

29. Su GL, Dorko K, Strom SC, Nüssler AK, Wang SC. CD14 expression and production by human hepatocytes. J Hepatol. 1999;31(3):435–442. doi:10.1016/S0168-8278(99)80034-8

30. Ancuta P, Kamat A, Kunstman KJ, et al. Microbial translocation is associated with increased monocyte activation and dementia in AIDS patients. PloS One. 2008;3(6):e2516. doi:10.1371/journal.pone.0002516

31. Gannon P, Khan MZ, Kolson DL. Current understanding of HIV-associated neurocognitive disorders pathogenesis. Curr Opin Neurol. 2011;24(3):275–283. doi:10.1097/WCO.0b013e32834695fb

32. Muñoz-Nevárez LA, Imp BM, Eller MA, et al. Monocyte activation, HIV, and cognitive performance in East Africa. J Neurovirol. 2020;26(1):52–59. doi:10.1007/s13365-019-00794-3

33. Adler MW, Rogers TJ. Are chemokines the third major system in the brain? J Leukoc Biol. 2005;78(6):1204–1209. doi:10.1189/jlb.0405222

34. Gras G, Kaul M. Molecular mechanisms of neuroinvasion by monocytes-macrophages in HIV-1 infection. Retrovirology. 2010;7:30. doi:10.1186/1742-4690-7-30

35. Pelphrey KA, Carter EJ. BRAIN MECHANISMS FOR SOCIAL PERCEPTION: LESSONS FROM AUTISM AND TYPICAL DEVELOPMENT. Ann N Y Acad Sci. 2008;1145:283–299. doi:10.1196/annals.1416.007

36. Khandaker GM, Cousins L, Deakin J, Lennox BR, Yolken R, Jones PB. Inflammation and immunity in schizophrenia: implications for pathophysiology and treatment. Lancet Psychiatry. 2015;2(3):258–270. doi:10.1016/S2215-0366(14)00122-9

37. Park KM, Bowers WJ. Tumor Necrosis Factor-alpha Mediated Signaling in Neuronal Homeostasis and Dysfunction. Cell Signal. 2010;22(7):977–983. doi:10.1016/j.cellsig.2010.01.010

38. Boiko A, Mednova I, Kornetova E, Semke A, Bokhan N, Ivanova S. Cell Adhesion Molecules in Schizophrenia Patients with Metabolic Syndrome. Metabolites. 2023;13:376. doi:10.3390/metabo13030376

39. Meixensberger S, Kuzior H, Fiebich B, et al. Upregulation of sICAM-1 and sVCAM-1 Levels in the Cerebrospinal Fluid of Patients with Schizophrenia Spectrum Disorders. Diagnostics. 2021;11:1134. doi:10.3390/diagnostics11071134

40. Yoon CY, Steffen LM, Gross MD, et al. Circulating Cellular Adhesion Molecules and Cognitive Function: The Coronary Artery Risk Development in Young Adults Study. Front Cardiovasc Med. 2017;4. Accessed April 7, 2023. https://www.frontiersin.org/articles/10.3389/fcvm.2017.00037

41. Mauliūtė M, Rugienė R, Žėkas V, Bagdonaitė L. Association of endothelin-1 and cell surface adhesion molecules levels in patients with systemic sclerosis. J Lab Med. 2020;44(6):343–347. doi:10.1515/labmed-2020-0050

42. Fabbri-Arrigoni FI, Clarke L, Wang G, et al. Levels of circulating endothelial cells and colony-forming units are influenced by age and dyslipidemia. Pediatr Res. 2012;72(3):299–304. doi:10.1038/pr.2012.76

43. Baysal A. Prognostic significance of sICAM-1 and sVCAM-1 molecules for cardiac surgery in pediatric patients with pulmonary hypertension. Anadolu Kardiyol Derg Anatol J Cardiol. Published online January 1, 2014. Accessed April 7, 2023. https://www.academia.edu/50755961/Prognostic_significance_of_sICAM_1_and_sVCAM_1_molecules_for_cardiac_surgery_in_pediatric_patients_with_pulmonary_hypertension

44. Lande MB, Adams HR, Kupferman JC, Hooper SR, Szilagyi PG, Batisky DL. A Multicenter Study of Neurocognition in Children with Hypertension: Methods, Challenges, and Solutions. J Am Soc Hypertens JASH. 2013;7(5):353–362. doi:10.1016/j.jash.2013.05.003

45. Lande MB, Batisky DL, Kupferman JC, et al. Neurocognitive Function in Children with Primary Hypertension. J Pediatr. 2017;180:148–155.e1. doi:10.1016/j.jpeds.2016.08.076

46. Tchalla AE, Wellenius GA, Sorond FA, et al. Elevated Soluble Vascular Cell Adhesion Molecule-1 Is Associated With Cerebrovascular Resistance and Cognitive Function. J Gerontol A Biol Sci Med Sci. 2017;72(4):560–566. doi:10.1093/gerona/glw099

47. Yong HYF, Batty NJ, Tottenham I, Koch M, Camara-Lemarroy CR. Soluble adhesion molecules: Cognitive worsening biomarkers in primary progressive multiple sclerosis? J Neuroimmunol. 2024;393:578384. doi:10.1016/j.jneuroim.2024.578384

48. Gregory MA, Manuel-Apolinar L, Sánchez-Garcia S, et al. Soluble Intercellular Adhesion Molecule-1 (sICAM-1) as a Biomarker of Vascular Cognitive Impairment in Older Adults. Dement Geriatr Cogn Disord. 2019;47(4-6):243–253. doi:10.1159/000500068

